# A cross-sectional analysis of the vaginal microenvironment in rheumatoid arthritis

**DOI:** 10.1101/2025.04.11.25325692

**Authors:** Marlyd E. Mejia, Savannah Bowman, Jessica Lee, Ali El-Halwagi, Keshia Ferguson, Maryjo Maliekel, Yixuan Zhou, Camille Serchejian, Clare M. Robertson, Mallory B. Ballard, Lee B. Lu, Sobia Khan, Olubunmi O. Oladunjoye, Shixia Huang, Sandeep K. Agarwal, Kathryn A. Patras

## Abstract

**Objective:** The human microbiota is implicated in the development and progression of rheumatoid arthritis (RA). Given the increased RA burden in women, and well-known correlations between the vaginal microbiota and local inflammation, we seek to understand the vaginal microenvironment in the context of RA pathology.

**Methods:** Self-collected vaginal swabs and questionnaires on dietary and health practices were obtained from 36 RA and 50 demographically-matched control women, 18-63 years of age. Additionally, medication regimen and disease activity and severity were captured for the RA cohort. Vaginal swabs were subjected to full-length 16S rRNA gene sequencing, multiplex cytokine analyses, and quantification of rheumatoid factor, c-reactive protein, and anti-citrullinated protein antibodies (ACPAs).

**Results:** Vaginal microbial richness and genera *Peptoniphilus* and *Prevotella*, among other rare taxa, were elevated in RA versus control samples. Vaginal IL-18 and EGF levels were increased in the RA group; IL-18 correlated with multiple microbial features whereas EGF levels were not associated with bacterial composition or other host factors. Within the RA cohort, decreased relative abundance of *Streptococcus* was associated with joint pathologies, and *Lactobacillus gasseri* was lower in individuals with serum detection of ACPAs and rheumatoid factor. Vaginal ACPAs were higher in the RA group and positively correlated with *Streptococcus* and multiple vaginal inflammatory cytokines.

**Conclusions:** We describe vaginal microbial and immunological differences in women with RA, particularly when accounting for diet and menopausal status, disease activity and severity, and medication use. This work opens a new avenue in the multidisciplinary approach to RA patient care.

## INTRODUCTION

Rheumatoid arthritis (RA) is an autoimmune disease affecting ∼1% of the global population(1, 2). RA manifests primarily as inflammatory arthritis characterized by synovial inflammation leading to permanent joint damage and decreased quality of life. RA is diagnosed based on the presence of joint pain and synovitis. The presence of anti-citrullinated protein antibodies (ACPA) and rheumatoid factor (RF) aids in the diagnosis. RA disease activity is followed using disease activity measures such as the clinical disease activity index (CDAI) which incorporates patient and physician overall assessment of disease activity and the number of swollen and tender joint counts. Severity and damage from RA is evaluated on radiographs, assessing joint space narrowing (JSN) and radiographic erosions (RE)(3). RA-related inflammation also has systemic implications and multi-organ involvement including elevated risk for cardiac disease, sleep disturbance, cognitive decline, and mucosal inflammation(4). These wide-reaching implications have highlighted the need for more integrated and multidisciplinary approaches to RA patient care.

Like other autoimmune diseases, RA is caused by a combination of underlying genetic factors and environmental triggers, including infection or microbial perturbations(5–7). Experimentally, transfer of fecal microbes from RA patients to arthritis-prone mice drive more severe synovial inflammation suggesting a causal role for the microbiome in RA pathology(8, 9). Microbial molecular mimicry, bystander activation, epitope spreading, or production of cryptic antigens (i.e., post-translational citrullination via microbial enzymes) can trigger an immune response cascading into auto-reactivity(10, 11). For example, release of citrullinated oral bacteria into circulation during periodontal disease can activate ACPA-producing B cells and correlates with RA symptom flares(12). Unique oral and gastrointestinal microbial signatures, such as increased *Prevotella* sp., are observed in new onset RA patients(13, 14), indicating RA in turn may influence microbial composition, even in the absence of mucosal disease. These studies strongly implicate the microbiome in the pathogenesis of RA and may have important clinical implications for patients.

RA has a predilection to affect women (∼70% of disease burden), with reproductive-age women having 3-5-fold increased RA incidence compared to male counterparts(1, 5, 15). Notably, hormonal contraception or pregnancy relieves symptoms in ∼75% of women, whereas disease aggravation is observed post-partum and post-menopause(16, 17). As with other mucosal sites, the vaginal microenvironment is impacted by environmental and host factors, being specifically sensitive to hormonal fluctuations during menses, throughout pregnancy, and in peri-menopause(18, 19). A non-optimal vaginal microbiota, typically characterized by low levels of *Lactobacillus* species, is associated with local or systemic inflammation, increased susceptibility to infections, and adverse gynecologic or obstetric outcomes(18, 20–22). Known associations between new onset RA or disease activity and microbial constituents, and the importance of vaginal microbe-immune interactions in women’s health, warrants characterization of the vaginal microenvironment in RA.

Hypothesizing that the vaginal microbiota and mucosal immunity are dysregulated in RA and correlate with autoimmune responses and disease activity, we performed microbiome, cytokine, and autoimmune factor profiling in vaginal samples collected from women with and without RA. We further integrated medical history, demographic information, medication use, and RA disease activity measures to resolve correlations with RA diagnosis and disease severity. We found that vaginal microbial and immunologic features are indicative of RA status and disease activity, revealing the vaginal mucosa as a previously unrecognized site impacted by RA.

## PATIENTS and METHODS

### Study patient demographics

Under written informed consent, we recruited women between 18-63 years of age from two outpatient rheumatology clinics in the Texas Medical Center, Houston, Texas: Baylor Medicine (“clinic 1”, May 2021 through February 2024) and Harris Health Smith Clinic (“clinic 2”, December 2023 through June 2024). The Institutional Review Board for Baylor College of Medicine (H-47537) and Harris Health System (23-05-3085) gave ethical approval for this work. Women diagnosed with seropositive and seronegative RA were included in the study. We used frequency matching on menopausal status, age (+/-5 years), educational status, and type 2 diabetes status to identify controls, with all four factors aligning between most individual comparisons (**Table 1**). Exclusion criteria consisted of active herpes lesions in the vulvo-vaginal area, an original diagnosis of juvenile idiopathic arthritis, active pregnancy, a history of medical problems that would otherwise impact sample collection, known HIV positivity, and antibiotic use within the last month. Eight RA and four controls did not disclose antibiotic use but were retained in the study.

**Table 1.**
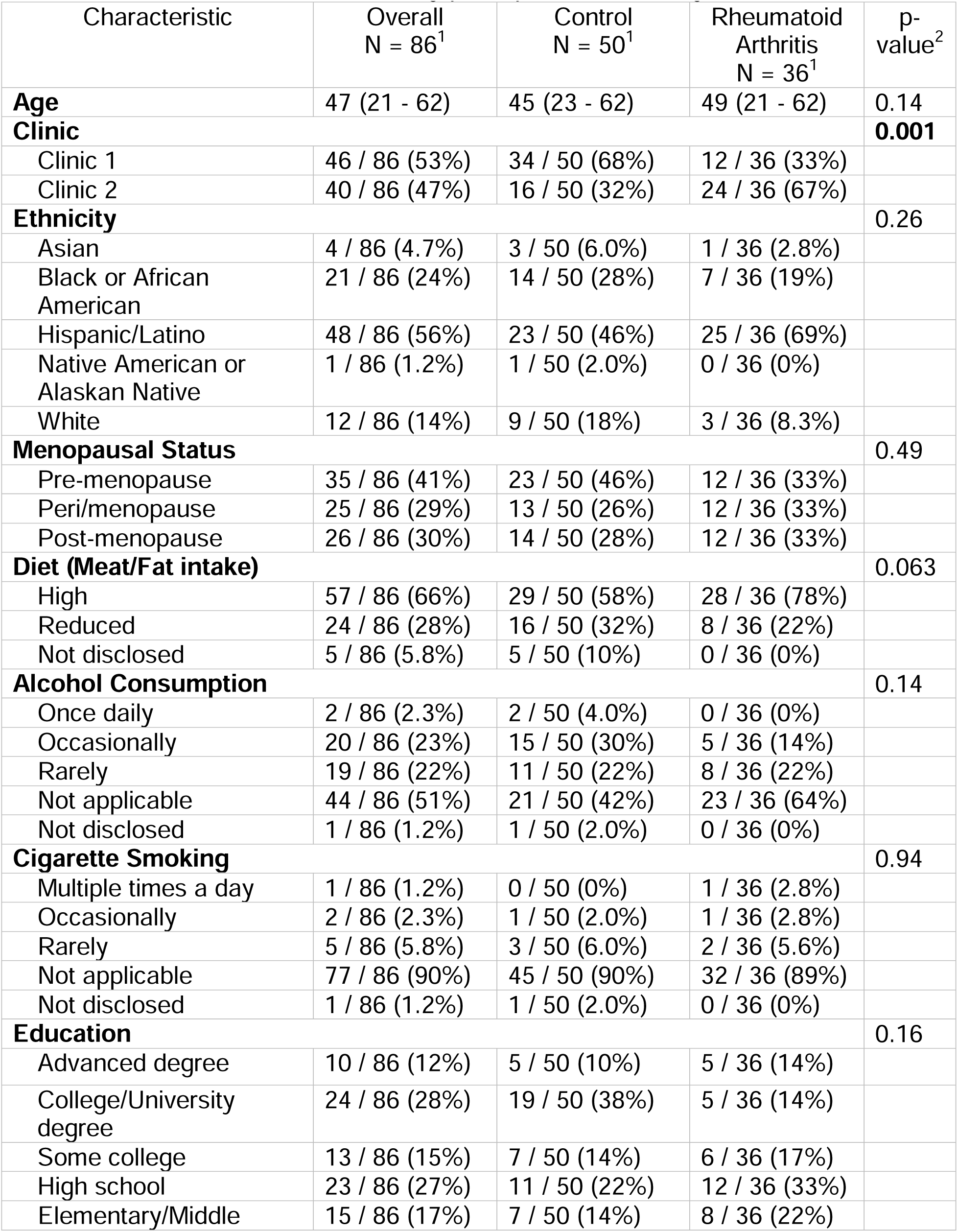
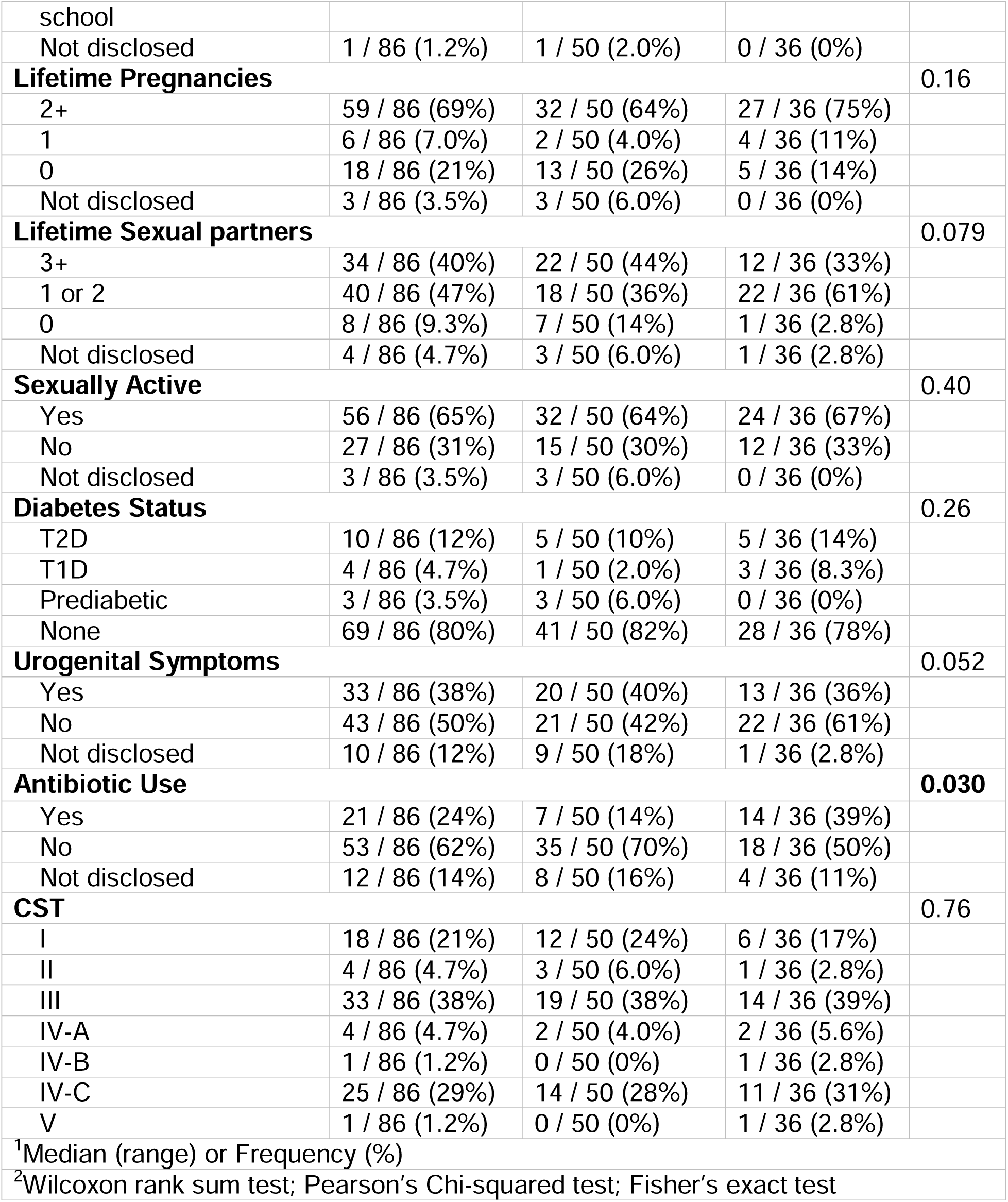
Clinical demographics of study participants stratified by RA status.

### Clinical Sample Collection

Subjects completed a questionnaire for self-reported dietary and health practices, provided in English or Spanish. For the RA cohort, clinicians reported current and prior medication use, radiographic and serological history, current CDAI score, and known gynecologic symptoms. Vaginal swabs were self-collected by patients instructed to rotate the swab four times clockwise and four times counterclockwise along the vaginal wall, repeating the process for a total of two swabs. Some control patient swabs were obtained by a clinician during routine procedures. Swabs were transported to the lab in a chilled anaerobic container, resuspended in 2mL of sterile phosphate buffered saline (PBS), and stored at-20°C.

### DNA extraction, 16S rRNA sequencing, and analyses

DNA extractions were performed using a quick-DNAFungal/Bacterial Microprep Kit (Zymo Research) per manufacturer protocol except for a 15-minute bead-beating step and elution in 40 µL molecular grade water. Long-read 16S rRNA genes were amplified via a 35-cycle PCR with barcoded 27F and 1492R primers and the HotStarTaq Plus Master Mix Kit (Qiagen) and purified using Ampure PB beads (Pacific Biosciences). Sample libraries were prepped using SMRTbell libraries (Pacific Biosciences) and sequenced by MR DNA Lab (Shallowater, TX, USA) on the PacBio Sequel using the diversity assay bTEFAP® LONG HIFI 5k following the manufacturer’s guidelines. Secondary analysis consisted of Circular Consensus Sequencing, using PacBio’s CCS algorithm. Barcodes were removed prior to downstream analysis.

R package Decontam (23)(R v4.4.2 (2024-10-31) -- “Pile of Leaves”) identified 12 Feature IDs for removal at a threshold of 0.2. Contaminant *Pseudomonas veronii* was manually removed. Reads were denoised using DADA2 with parameters--p-max-ee 2,--p-trunc-q 2,--p-min-len 1000, and--p-max-len 1600 using QIIME2 v2023.5(24). Operational Taxonomic Units (OTUs) were assigned using the Greengenes2 reference tree(25). Alpha diversity (OTUs and Shannon), beta diversity (weighted normalized unifrac distance and PERMANOVA), and differential abundance (ANCOM) tests were carried out in QIIME2(26). Output files were exported and analyzed in R Studio v2024.12.0+ using packages phyloseq v1.50.0(27), gtsummary v2.1.0, dplyr v1.1.4, plyr v1.8.9, ggpubr v0.6.0, lmtest v0.9.40, tidyverse v2.0.0, car v3.1.3, boot v1.3.31, pROC v1.18.5, and nlme v3.1.168. Data visualization was performed using ggplot2(28) and GraphPad Prism v10.2.3 (GraphPad Software, Inc.). For linear and logistic regression models, only species that were present in >6% of samples were assessed for correlation. In the RA group only, taxa that were present in at least 3 samples were prioritized for visualization.

### Cytokine quantification and analyses

Vaginal samples underwent a maximum of three freeze-thaw cycles prior to cytokine analyses. Tubes were centrifuged at 10,000 rcf for 15 minutes prior to being run on a 48-plex MILLIPLEX® Human Cytokine/Chemokine/Growth Factor Panel A Magnetic Bead Panel (Cat#: HCYTA-60K-PX48). PBS was used as the matrix solution for standards and controls following the company’s instructions. The instrument passed calibration using the Bio-Plex Calibration Kit (Cat#: 171-203060) and passed validation using the Bio-Plex Validation Kit 4.0 (Cat#: 171-203001).

A Logistic-5PL regression type was used to generate analyte standard curves. Cytokines with a zero or negative FI-background value were deemed out of range and assigned a concentration of 0.001. Of the 48 cytokines tested, 38 were kept for analyses. GRO-α was excluded because of incongruities between runs. No normalization was conducted as all other concentration ranges were comparable between runs, and total protein in vaginal fluid varies widely(29). Fractalkine, IL-3, IL-7, IL-13, IL-17A, IL17-E/IL-25, IL-17F, MIP-1α, and PDGFF-AB/BB were undetected in the majority of samples and were excluded from analyses.

### Statistics

Single timepoint vaginal swabs are represented by individual symbols on all plots. Contingency and frequency data were analyzed by Fisher’s Exact test. Microbial, cytokine, or protein comparisons were analyzed by two-tailed Mann-Whitney U test. Differential taxa were identified via ANCOM. Two-tailed Spearman correlations were performed between microbial and metadata factors and between microbes and immune factors. Multiple linear, non-linear and logistic regression models incorporated variables that were significantly different in groups (i.e., antibiotic use and lifetime sexual partners). Menopausal stage and dietary groups were always included, and serum ACPA detection added to within-RA analyses (except with RF and CDAI because of collinearity). Bacteria included in the logistic regression model (**Fig. 1**) were determined using the backwards deduction method. Cytokine and ACPA-pertaining models included stepwise addition of significant variables, limiting the number of variables, and choosing the best model based on lower AIC. Coefficients and *P*-values for significant regression model variables are provided in **Table S1**. Variable coefficients and adjusted *P*-values are shown unless otherwise stated. Mann-Whitney U test *P*-values are shown for comparisons significantly identified by regression analysis. Kruskal-Wallis with Dunn’s multiple comparisons test was used in comparisons with positive or negative detection of morphologic or serum markers. All data were considered non-parametric and non-paired. *P*-values <0.05 were considered statistically significant.

**Figure 1.**
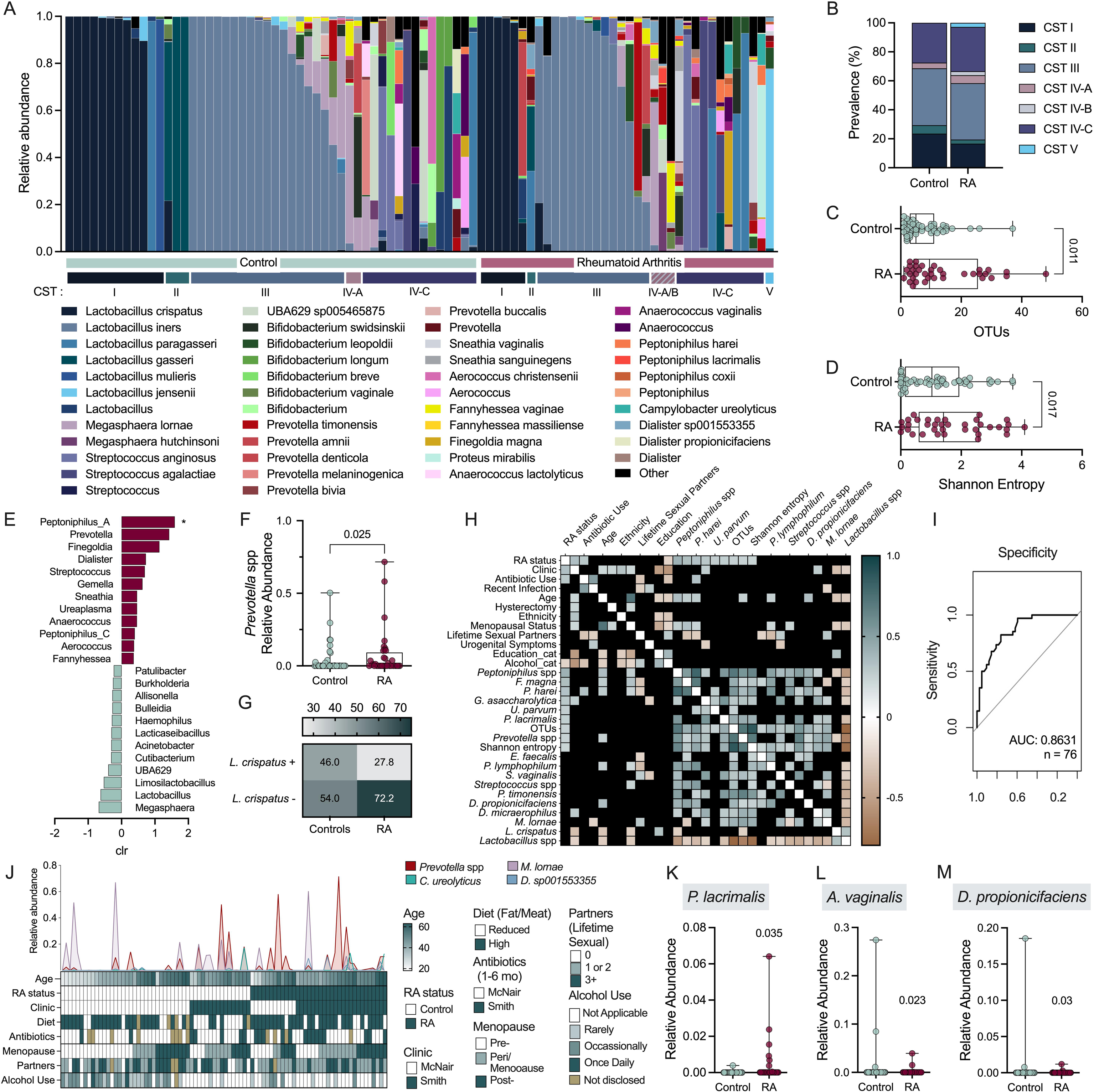
Low abundance taxa distinguish the vaginal microbiota of those with and without RA. Vaginal samples of RA (*n*=36) and controls (*n*=50) were subjected to long-read 16S rRNA gene sequencing. (**A**) Relative abundances of the top 15 genera, broken down by species, and their respective community state type (CST). (**B**) CST prevalence in each group. Alpha diversity measured by (**C**) OTUs and (**D**) Shannon entropy. (**E**) ANCOM-determined genera associated with disease state. (**F**) *Prevotella* spp. abundance and (**G**) *L. crispatus* presence between groups. (**H**) Clinical and microbial variables most strongly correlated with RA (unadjusted, non-significant values blacked out). (**I**) ROC curve of logistic regression model predicting RA as an outcome of (**J**) variables included in current and future regression analyses and the relative abundances of microbes used in logistic regression; (I) excludes lifetime sexual partners. Distribution of relative abundances of (**K**) *Peptoniphilus lacrimalis*, (**L**) *Anaerococcus vaginalis* and (**M**) *Dialister propionicifaciens* in each group. (A) Bars and (C,D,F,K-M) symbols represent individual subjects. Data were analyzed by (B,G) Fischer’s exact test, (C,D,F) Mann-Whitney U test, (E) ANCOM (significance denoted by *), (H) unadjusted Spearman correlation, multiple logistic regression (I) AUC and (K-L) predictors of RA. Significant *P*-values are shown.

## RESULTS

### Patient demographics and RA disease heterogeneity

Female RA patients (*n* = 36) and healthy controls (*n* = 50) self-collected vaginal swabs. Demographics including age, race and ethnicity, menopausal status, educational status, alcohol and cigarette use, sexual activity, and type 2 diabetes status were similar between groups whereas antibiotic use within the last 6 months was greater in the RA group (**Table 1**). Additional medical conditions including additional autoimmune diseases, current medications and urogenital symptoms, and recent infections are recorded in **Fig. S1**. The RA cohort displayed heterogeneity with a time since diagnosis ranging from less than one to 22 years, symptoms ranging from high disease activity to remission, and varied current and prior medication history (**Fig. S2**).

### Alpha diversity and lesser abundant taxa differentiate RA from control vaginal microbiota

Vaginal microbiomes are grouped into community state types (CSTs) based on the presence of one of four primary *Lactobacillus* species (*L. crispatus*, *L. gasseri*, *L. iners*, and *L. jensenii* as CST I, II, III and V, respectively)(20, 30). In about 25% of women, a non-optimal vaginal microbiome, characterized by a mixture of anaerobic organisms including *Gardnerella* or *Prevotella* spp. (CST IV), is associated with increased risk for adverse gynecological outcomes(20, 30). To determine whether women with RA had altered vaginal microbiomes, 16S rRNA gene sequenced vaginal compositions were categorized into CSTs using the VALENCIA classifier(31)(**Fig. 1A**). CST prevalence was comparable between RA and controls (**Fig. 1B**). In terms of alpha diversity, the RA cohort displayed elevated OTUs (**Fig. 1C**) and greater richness and evenness as measured by Shannon Entropy (**Fig. 1D**). Analysis of compositions of microbiomes (ANCOM) identified differential relative abundant genera with *Peptoniphilus*, *Prevotella*, *Finegoldia*, and *Dialister* associated with RA, and *Megasphaera*, *Lactobacillus* and *Limosilactobacillus* associated with controls (**Fig. 1E**). The *Prevotella* signature was retained in the RA group when assessed by Mann-Whitney U-test (**Fig. 1F**). Although *L*. *crispatus* was more likely to be detected in the control versus RA group with an odds ratio of 2.2, this difference was non-significant (95% CI: 0.87-5.5; *p=*0.067, **Fig. 1G**).

Spearman correlation showed that RA status was positively associated with clinic (site of recruitment), antibiotic use, and microbial features including *Peptoniphilus* (and *P. harei* and *P. lacrimalis*), *Finegoldia magna, Gemmela assacharolytica, Ureaplasma parvum,* and *Prevotella* (**Fig. 1K**). We further performed logistic (clinical variables) and linear (microbial relative abundances) regression analyses (**Fig. 1H**) to develop a co-variate adjusted receiver operating characteristic (ROC) curve using backwards selection of input variables to predict RA status (**Fig. 1I**). We incorporated clinic site, diet, antibiotic use, menopausal status, and alcohol use, along with relative abundance of *Prevotella*, *Dialister sp001553355, Megasphaera lornae,* and *Campylobacter ureolyticus,* which generated a ROC area under curve (AUC) of 0.86 (**Fig. 1I,J**). Variables incorporated in adjusted regression models in the ROC curve and subsequent analyses are shown in **Fig. 1J** and **Table S1**. Multiple linear regressions showed that, individually, *P. lacrimalis* relative abundance increased in the RA group (**Fig. 1K**) whereas *Anaerococcus vaginalis, and Dialister propionicifaciens* were increased in the controls (**Fig. 1L-M**).

### RA clinical factors correlate with vaginal microbial signatures

Oral and gastrointestinal microbial signatures are correlated with RA disease activity and symptom flares(12, 32). While bacteria can modulate inflammation directly, microbial communities are also responsive to inflammatory environments(33, 34). To determine whether RA is likewise associated with vaginal microbial composition, we assessed correlations between radiographic changes from RA or autoantibody presence with vaginal microbes. Joint space narrowing (JSN) corresponds to cartilage damage and radiographic erosions (RE) to bone destruction (3). RA patients with documented JSN or RE had phylogenetically distinct vaginal compositions compared to controls, whereas RA patients without erosive disease were comparable to controls (**Fig. 2A-B**). No compositional differences were seen between controls and RA patients with and without detection of serum ACPA and RF (**Fig. 2C-D**). Multiple logistic regression determining bacterial abundance as an outcome of erosion or seropositivity within the RA group was used to detect taxonomic differences (controls depicted for comparison). *Streptococcus* relative abundance was decreased in JSN+ individuals with a non-significant reduction in RE+ individuals (**Fig. 2E-F**). Despite overlapping phylogenetic compositions, both ACPA+ and RF+ groups showed reduced relative abundance and prevalence of *L. gasseri* compared to negative-testing counterparts, and RF+ status was also associated with lower *P. harei* and *P. colorans* (**Fig. 2G-H**).

**Figure 2.**
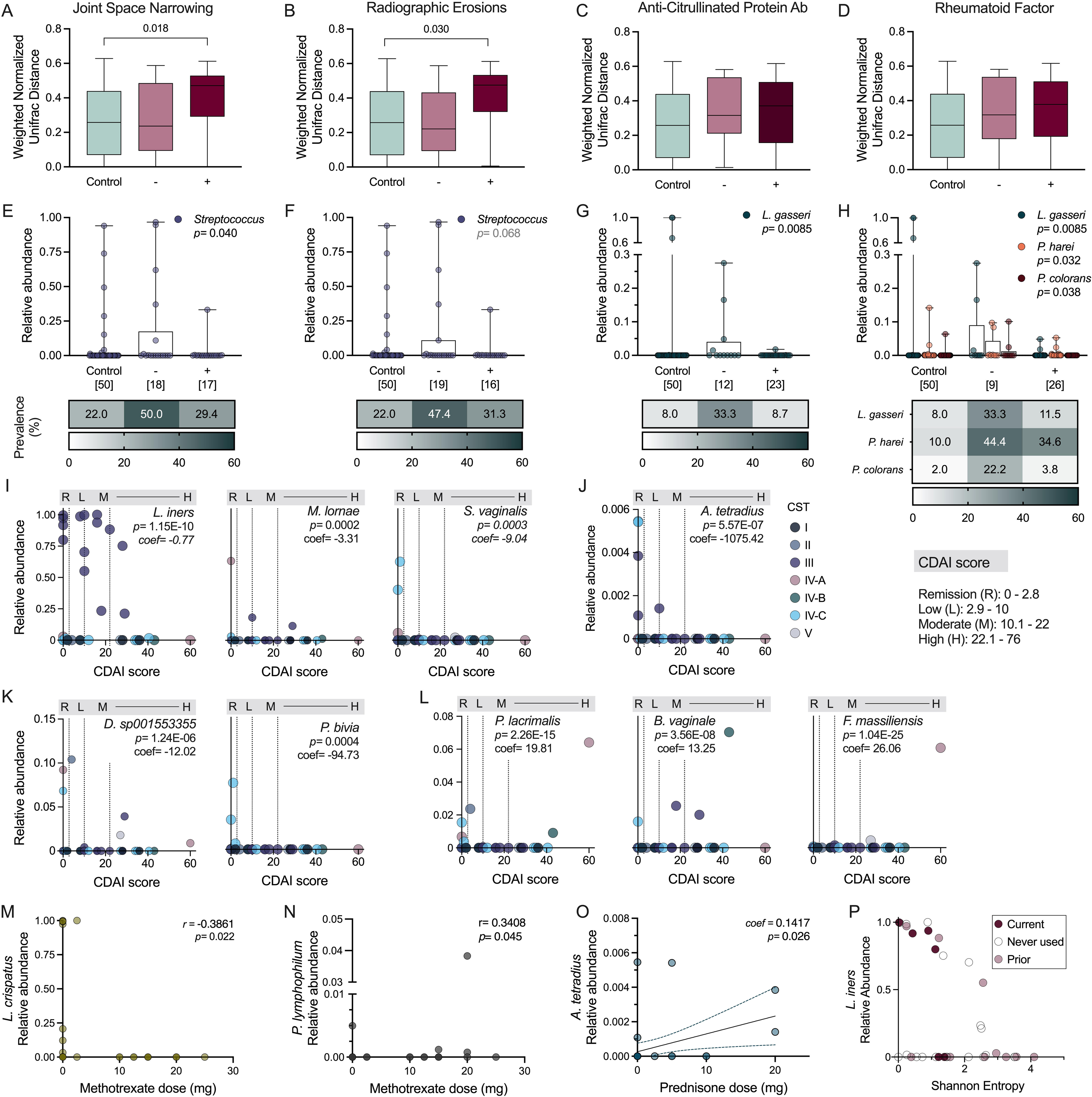
Vaginal microbial signatures differ across RA disease severity metrics. Weighted normalized unifrac distances with binary outcome variables (negative or positive detection) for (**A**) JSN, (**B**) RE, (**C**) serum ACPA, and (**D**) serum RF detection, and compared to controls. Relative abundances and prevalence of taxa associated with (**E**) JSN, (**F**) RE, (**G**) serum ACPA, and (**H**) serum RF as informed by logistic regression analysis with controls for comparison. (**I**) Highly abundant, (**J**) least abundant, and (**K**) low abundant taxa inversely associated with CDAI score, and (**L**) low abundant taxa positively associated with CDAI score. Dose association between methotrexate and relative abundance of (**M**) *L. crispatus* and (**N**) *P. lymphophilium* and prednisone with (**O**) *A. tetradius.* (**P**) Relative abundance of *L. iners* as an interaction between Sulfasalazine and Shannon entropy. (A-D) Tukey’s box and whisker plots. (E-P) Symbols represent individual subjects. Prior medication usage (P) was combined with non-use and analyzed by multiple logistic regression against current use. Data were analyzed by (A-D) PERMANOVA comparing controls and RA subgroups, (E-H,P) multiple logistic regression determining bacterial abundance as an outcome of positive/negative or non/current user status within the RA group, and (I-O) multiple linear regression. Significant adjusted *P*-values are shown.

The Clinical Disease Activity Index (CDAI) score reflects the current activity of disease at the time of sample collection (35, 36). Using multiple linear regression, we observed inverse correlations with both higher abundance (*L. iners, M. lornae,* and *S. vaginalis*), and lower abundance (*A. tetradius* and *Dialister sp001553355*) taxa and CDAI score ranges (**Fig. 2I-K**). *P. bivia* was only detected in patients during remission (**Fig. 2L**). Conversely, *P. lacrimalis*, *F. massiliensis* and *B. vaginale* were positively correlated with CDAI, although these taxa were present in only a subset of RA patients (**Fig. 2L**).

To determine whether disease modifying anti-rheumatic drugs (DMARDs) and biologics impact vaginal microbial composition, as seen in the gut(37), we performed multiple linear regression of current medication dosage and microbial relative abundances. Methotrexate dose, the most common medication in our cohort (**Fig. S1**), was inversely correlated with *L. crispatus* and positively correlated with *P. lymphophilium* (**Fig. 2M-N**). Increased prednisone dose was positively correlated with rare taxa *A. tetradius* (**Fig. 2O**). Sulfasalazine was positively correlated with relative abundance of *L. iners,* a phenomenon that was more pronounced when considering the interaction between the medication and overall composition (**Fig. 2P**).

### Specific vaginal cytokines are elevated in RA and levels are influenced by diet and menopausal status

Microbial signatures corresponding with RA could potentially cause, or derive from, local inflammation(33, 38). To correlate vaginal microbes and local immune mediators, we performed 48-plex cytokine analyses of vaginal samples. Of 38 detected cytokines, interleukin (IL)-18, epidermal growth factor (EGF), and tumor necrosis factor (TNF), were significantly higher in RA samples, with a non-significant increase in IL-12p(70) and no variation seen in other cytokines including RA-associated IL-1β, IL-6, or IL-15 (**Fig. 3A-G**). RA status alone did not explain the total variance among vaginal immune profiles (**Fig. 3H**). While fewer lifetime sexual partners were recorded in the RA group (**Table 1**), this did not contribute to vaginal cytokine variability across groups (**Fig. 3I**). Conversely, and as with microbial analyses, we observed impacts of diet and menopausal status on cytokine distributions in the context of RA (**Fig. 3J-K**). When stratified by diet (fat/meat intake), sCD40L, eotaxin, IL-8, IL-10, and IL-27 were increased in RA patients compared to controls in individuals with reduced fat/meat consumption (**Fig. 3L**). When stratified by menopausal status, postmenopausal RA patients showed elevated granulocyte-macrophage colony-stimulating factor (GM-CSF), IL-2, IL-12(p40), and transforming growth factor (TGF), compared to post-menopausal controls (**Fig. 3M**). When stratified by both diet and menopausal status, additional cytokines interferon (IFN)γ, IL-1α, IL-9, and IL-22 were significantly elevated in post-menopausal, reduced fat/meat diet RA patients compared to matched controls (**Fig. 3N**). Similarly, IFNγ was elevated in peri/menopausal RA patients on reduced fat/meat diet compared to controls (**Fig. 3N**). Pre-menopausal RA patients on a high fat/meat diet displayed decreased IL-1α and IL-9 compared to controls (**Fig. 3N**). Elevated IL-18 concentrations corresponded with RA-associated microbial signatures of *Prevotella*, *Peptoniphilus*, *A. vaginalis*, *C. ureolyticus*, and increased OTUs via Spearman correlation. TNF was also associated with *Prevotella* whereas *Peptoniphilus* correlated with IL1-α, OTUs were correlated with IFNα2, and both *D*. sp01553355 and *P. lacrimalis* were inversely correlated with six immunological markers (**Fig. S3A-B**). Prediction of RA using bootstrapping of a logistic regression model combining *Prevotella*, *Peptoniphilus*, EGF, the interaction between sCD40L and diet, menopausal status and clinic resulted in a mean AUC of 0.97 comparing 34 RA to a subset of 13 controls and 0.93 compared all 35 RA to the remaining 22 controls (95% CI [0.90, 1] and CI [86, 99], respectively) (**Fig. S3C-F**).

**Figure 3.**
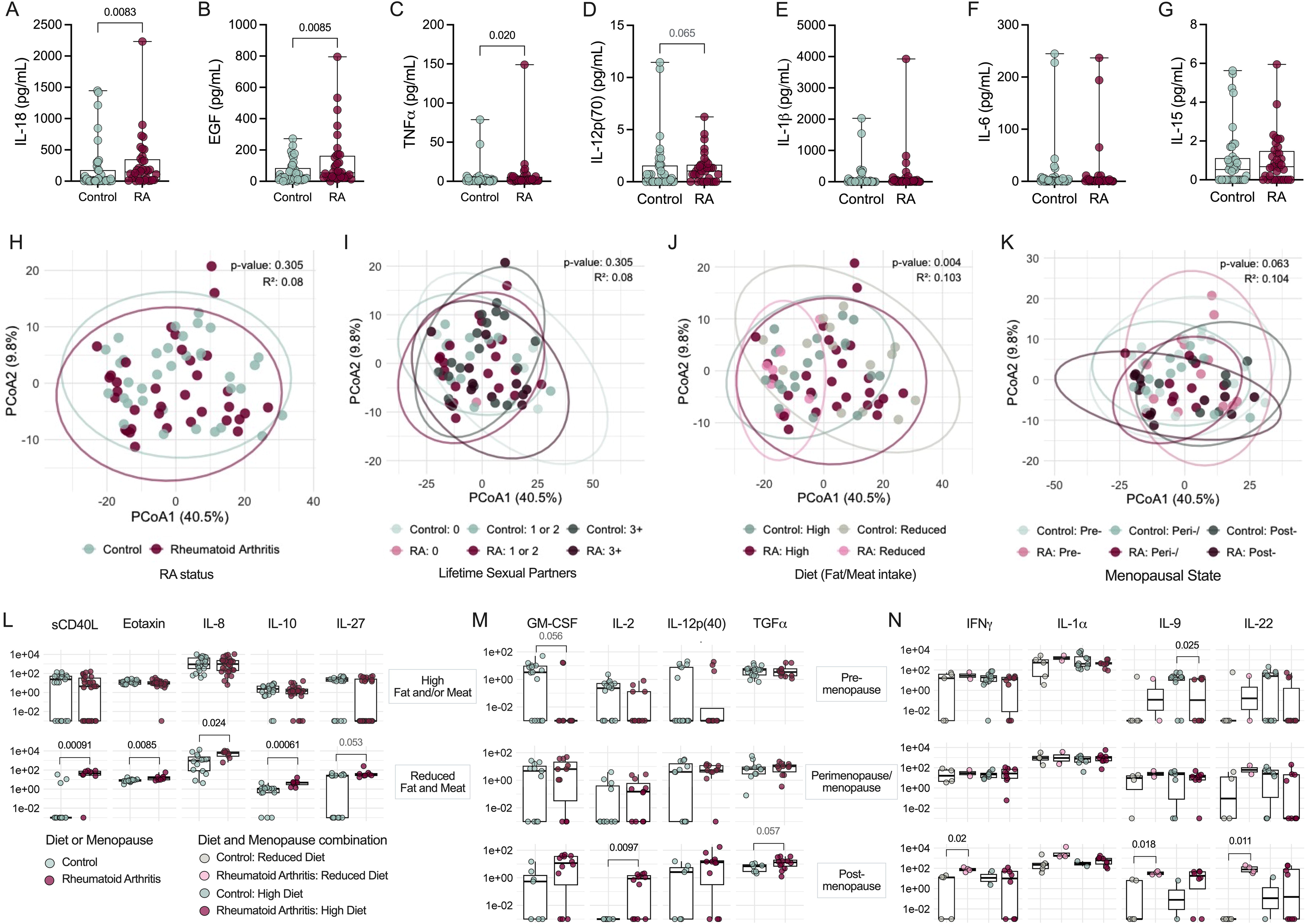
RA is associated with increased vaginal inflammation when stratified by menopausal and dietary factors. Vaginal swab samples were analyzed using a 48-plex ELISA. Unadjusted comparisons of differential (**A**) IL-18, (**B**) EGF, (**C**) TNF, (**D**) IL-12p(70), and non-differential (**E**) IL-1β, (**F**) IL-6, and (**G**) IL-15 in vaginal fluid of control and RA subjects. Overall immune profiles clustered by (**H**) RA status and RA-stratified (**I**) history of lifetime sexual partners, (**J**) diet described by either high fat and/or meat intake (high) or both low fat and meat intake (low), and (**K**) menopausal status. Immunological markers differentially expressed in the RA group when stratifying by (**L**) diet, (**M**) menopausal status, and a combination of (**N**) diet and menopausal status. Symbols represent individual subjects. Data were statistically analyzed by (A-D, L-N) Mann-Whitney U test, (H-K) PERMANOVA. (L-N) Multi-linear regression analysis shown in **Table S1**. Significant *P*-values are shown.

### Vaginal cytokine levels are correlated with RA clinical factors

Multiple studies demonstrate elevated circulating inflammatory cytokines in RA patients with active disease versus those in remission/quiescence(39, 40). To assess whether vaginal cytokine signatures correspond with disease metrics, we performed adjusted logistic regression analyses within the RA cohort. Vaginal macrophage-derived chemokine (MDC) levels were decreased in women with JSN (**Fig. 4A**), whereas detection of serum RF was associated with decreased EGF, lymphotoxin (LT)-α, and monocyte chemotactic protein (MCP)-3 (**Fig. 4B-D**). No associations with radiographic erosions nor serum ACPA and vaginal cytokines were observed. When assessing CDAI score and vaginal cytokine associations, interestingly, IL-18 was the only cytokine inversely correlated with disease activity (**Fig. 4E**). Thirty-one additional cytokines were positively correlated with CDAI using a Poisson distribution model adjusted for diet and menopausal status. Sixteen cytokines were related to RA diagnosis in association with diet, menopausal status, or both (**Fig. 3**), except for EGF, IL-8, IL-9, and IL-22 (**Fig. 4E**). The other sixteen cytokines were uniquely linked to CDAI (**Fig. 4E**). *D*. sp01553355 and *P. lacrimalis* negatively correlated with CDAI score and 14 CDAI-associated immune markers. *P. bivia*, *M. lornae*, *B. vaginalis*, and *F. massiliensis* correlated with immune markers that increased with CDAI score except *M. lornae* and MDC (**Fig. S3A-B**).

**Figure 4.**
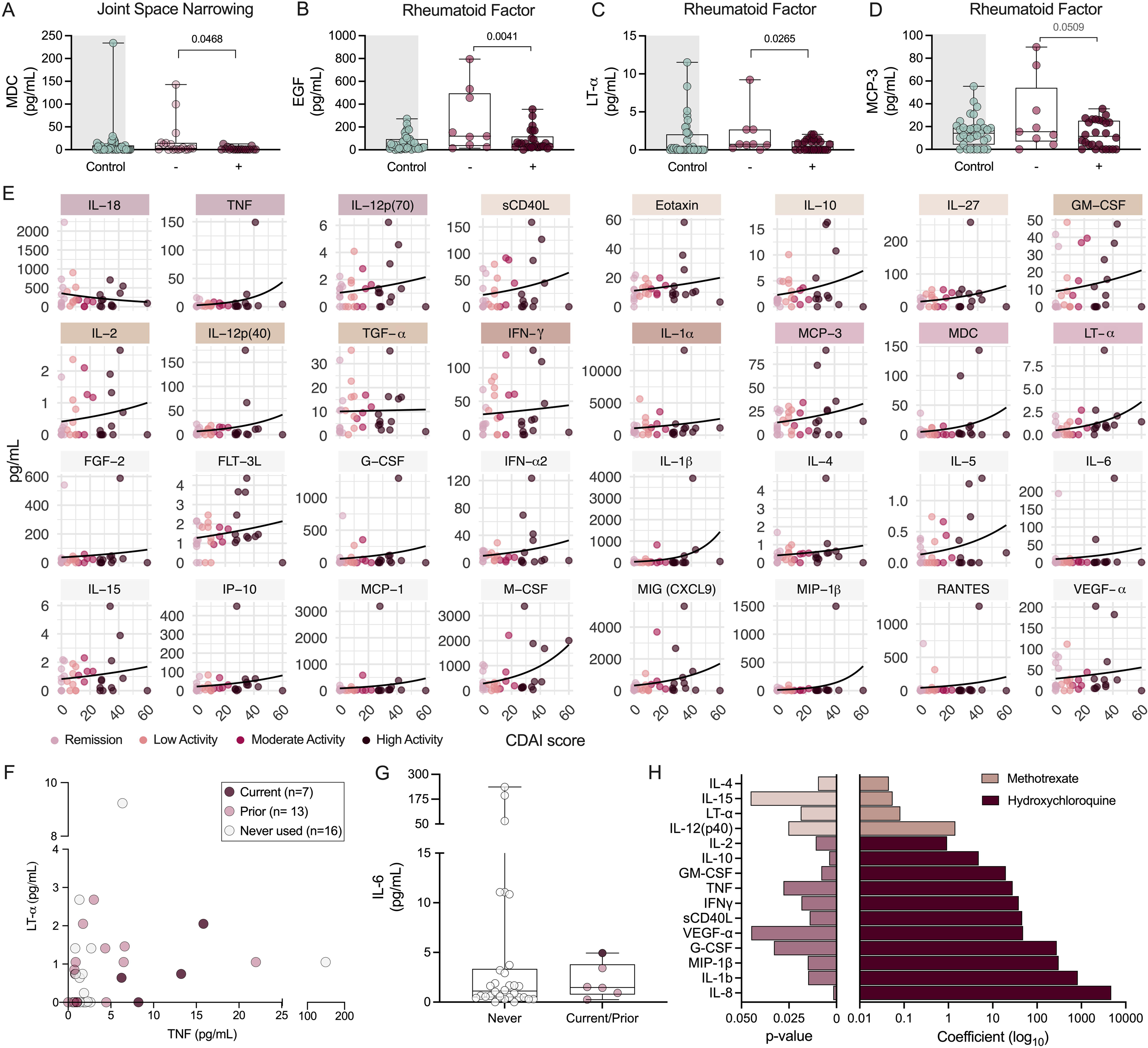
Elevated inflammatory markers correlate with increased CDAI score but not morphologic or serologic RA manifestations. Distributions of (**A**) MDC based on JSN status and (**B**) EGF, (**C**) LT-α, and (**D**) MCP-3 based on serum RF positivity status. Controls are depicted as a reference. (**E**) Cytokines predictive of CDAI score when accounting for diet and menopausal bins. Use of (**F**) TNF and (**G**) IL-6 inhibitors and their corresponding concentrations in vaginal samples. (**H**) *P*-values and coefficients of methotrexate dose and hydroxychloroquine use in predicting cytokine concentration. (A-G) Symbols represent individual subjects. Immune factors colored by RA association: control vs RA, diet subgroups, menopausal subgroup, diet: menopausal subgroup, and JSN/RF detection. Data were analyzed by (A-D) logistic regression for positive and negative detection of factors within the RA group, accounting for diet and menopausal status. (E) Non-linear regression using non-adjusted Poisson distribution with 95% CI. (F-G) Multiple logistic regression predicting current medication use as a factor of cytokine concentrations. Prior use colored for reference but combined with (F) never used or (G) current group. (H) Multi-linear model predicting cytokine as an outcome of medication use when adjusted for menopausal status, clinic, and diet. Multi-linear regression analysis shown in **Table S1**. Significant *P*-values shown for (A-D, F-H).

To observe whether biologics impact vaginal cytokines, we assessed anti-TNF (Adalimumab, Certolizumab_pegol, Etanercept, Golimumab and Infliximab) and anti-IL-6 (Tocilizumab and Sarilumab) medication use and their corresponding vaginal cytokines. Vaginal TNF and LT-α (a TNF family cytokine that binds the same receptor) concentrations did not differentiate patients currently taking anti-TNF medication from those with prior or no history of use (**Fig. 4F**). Due to small numbers, current and prior anti-IL-6 users were assessed together against non-users, and no medication-mediated vaginal IL-6 suppression was detected (**Fig. 4G**). Women taking methotrexate exhibited slight, but significantly increased, IL-4, IL-12(p40), IL-15, and LT-α (**Fig. 4H**). Hydroxychloroquine use was associated with increased levels of eleven cytokines. No statistical significance was found between IL-18 and medication use (Fig. S3G-H). No correlations were found between microbes and cytokines associated with medication use.

### Vaginal ACPA predominantly correlates with *Streptococcus* and inflammatory signatures in RA

Serum ACPA and RF informs RA diagnosis while CRP captures inflammation; however, the latter two are non-RA-specific(41–43) and all three factors can be induced by microbial-mediated inflammation(41, 44). We quantified vaginal ACPA, RF, and CRP levels via ELISA (**Fig. 5A-C**). ACPA was elevated in RA patients with no differences observed in RF levels (**Fig. 5A-B**). ACPA and RF were positively correlated in the RA group but not controls (**Fig. 5D**). ACPA was detected at higher frequency within the RA cohort (OR=6.750) while RF could be detected in about half of all women (**Fig. 5E**). Within the RA group, serum and vaginal detection of ACPA or RF were not interrelated (**Fig. 5F**), limiting the potential of vaginal levels to serve as a proxy for serum ACPA and RF(44). Within the RA group, ACPA and RF positively correlated with *Streptococcus* and *L. crispatus*, respectively (**Fig. 5G-H**). IL-15 was positively correlated with ACPA levels in both groups (**Fig. 5I**). Within the RA group, twenty-one additional cytokines were positively correlated with vaginal ACPA levels, ten of which positively correlated with *Streptococcus* abundance (**Fig. 5J**, **Fig. S3A-B**, **Fig. S4A**). Several of these had overlapping positive correlations with RF including IL-6, RANTES, FGF-2, and G-CSF, with positive correlations for FGF-2 also observed in the control group (**Fig. 5J, Fig. S4A-B**). Neither ACPA nor RF were correlated with CDAI despite shared associated cytokines (**Fig. 5K-L**).

**Figure 5.**
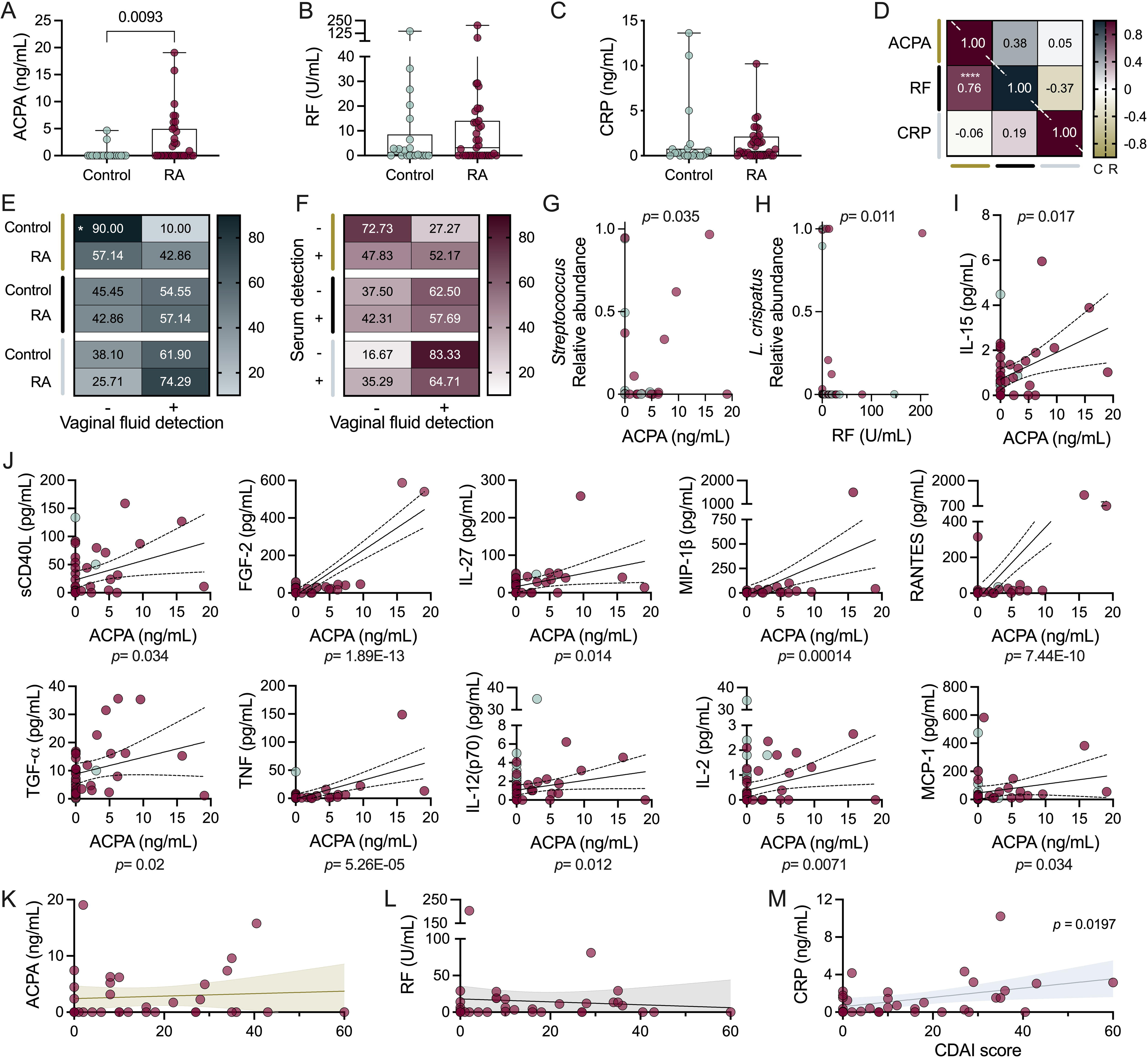
RA biomarkers are detected in the vaginal tract and ACPA levels correlate with elevated inflammatory cytokines predominantly in the RA cohort. Vaginal concentrations of (**A**) ACPA, (**B**) RF, and (**C**) CRP. (**D**) Correlated detection of vaginal ACPA, RF, or CRP in controls (“C”, upper right) and RA cohort (“R”, bottom left). (**E**) Prevalence of ACPA, RF, or CRP in control and RA vaginal samples. Within individuals of the RA cohort, (**F**) co-detection of serum and vaginal ACPA, RF, and CRP. Microbial correlations with biomarkers: (**G**) ACPA with *Streptococcus* spp and (H) RF with *L. crispatus*. Correlations to ACPA levels for cytokines also associated with *Streptococcus* abundance in (**I**) both control and RA and (**J**) only RA groups. (**K**) ACPA, (**L**) RF, and (**M**) CRP level correlations with CDAI score. Symbols represent individual subjects. (G-M) Spearman correlations and 95% CI are shown for immune markers found significant in multiple linear regression. Data were analyzed by (A-C) Mann-Whitney U test, (D) Spearman correlation, (E-F) Fishers Exact test, (G-M) multiple linear regression. (A-C, G-M) Significant *P*-values shown, (D-F) \**P*=0.015, \*\*\*\**P*<0.001.

CRP was detected in the majority of RA and controls irrespective of genitourinary symptoms or vaginal ACPA and RF (**Fig. 5C-E, Fig. S4C-D**). Consistent with prior studies, vaginal CRP detection and concentrations did not reflect systemic CRP values(42, 43) (**Fig. 5F, Fig. S4E**). Microbially, CRP was associated with *L. paragasseri* relative abundance (**Fig. S4F**), In terms of RA disease activity, vaginal CRP positively correlated with CDAI, whereas vaginal ACPA and RF did not (**Fig. 5K-M**). Cytokines associated with vaginal CRP included negative correlations with IL-9 in both RA and control groups, and positive correlations with PDGF-AA and VEGF in the control group only (**Fig. S4F**).

## DISCUSSION

Rheumatoid arthritis is multisystem disease that can lead to irreversible joint damage and decreased quality of life. Despite microbial ties to RA(45), and clinical associations between RA, infertility, and adverse reproductive outcomes(46–48), the vaginal environment has not been described in the context of RA. Our study determined that the RA vaginal microenvironment is distinguished by higher alpha diversity, increased presence of taxa such as *Peptoniphilus*, and higher levels of growth factors and inflammatory cytokines particularly in postmenopausal stages and diet subsets. Most notably, ACPA was significantly higher in RA vaginal samples, and strongly correlated with *Streptococcus* and multiple inflammatory cytokines. Vaginal RF and ACPA levels highly correlated within the RA cohort, but not the controls, strengthening an association between these vaginal markers and RA. Yet, the discordance between vaginal and serum levels of these same factors suggests divergence between systemic and vaginal autoimmune features.

A recent study found that women with systemic lupus erythematosus (SLE) had vaginal microbiomes characterized by increased Shannon diversity and genera *Peptoniphilus* and *Streptococcus* compared to controls(49). Likewise, we found that women with RA had higher alpha diversity, *Peptoniphilus*, which has been linked to chronic wounds, osteoarticular infections, and bacterial vaginosis(50), and, in women with higher levels of vaginal ACPA, *Streptococcus*. However, vaginal taxa associations are not consistent across autoimmune diseases: the RA cohort did not demonstrate significantly decreased *Lactobacillus* or increased *Bifidobacterium* reported in the SLE study. While RA is linked to rare gut and oral microbes absent in the vaginal tract(13, 14, 51), RA is consistently associated with *Prevotella* across mucosal sites, now including the vaginal tract. Vaginal *Prevotella* can be inflammatory and is frequent in bacterial vaginosis(38, 52). Within RA patients, women on higher methotrexate doses lacked *L. crispatus* and women on sulfasalazine were more likely to have higher *L. iners* abundance. Together, these taxonomic alterations suggest women with RA may have a higher risk of negative gynecologic outcomes.

Elevated vaginal EGF was a consistent RA signature irrespective of menopausal status, diet, microbial profile, CDAI score, or vaginal ACPA. EGF induces estrogen production and mediates estrogen-like effects to induce vaginal epithelial growth(53). Through engaging the EGF receptor, EGF downregulates TNF-induced apoptosis and necroptosis. This presents a dual role for EGF in promoting tissue regeneration and suppressing inflammation, while also driving problematic proliferation of synovial fibroblasts and osteoclasts in RA joints(54). Relatedly, reduced vaginal MDC in JSN+ individuals contrasts with elevated levels of MDC in damaged synovial joints(55). Vaginal IL-18, known to recruit neutrophils in response to infection(56), was associated with eight microbial features uniquely in RA, suggesting an important role in host-microbe dynamics. Although RA vaginal samples demonstrated elevated inflammation overall, these signatures were distinct from iconic, pathogenic inflammation in RA serum or synovial joints.

CDAI score, an assessment of current disease activity, yielded the most microbial and immunologic correlations within the RA cohort. However, increasing CDAI score was not correlated to vaginal ACPA, suggesting independent mechanisms for CDAI and ACPA-immune associations. ACPAs are generated in response to peptide citrullination via host or bacterial peptidylarginene deaminase (PAD) enzymes or citrullinated bacteria themselves(6, 7, 12). Vaginal autoantibodies correlate with bacterial vaginosis implying a connection to vaginal microbiota(57–59). Furthermore, mucosal ACPAs are site-specific, supporting the local origin of vaginally-detected ACPAs (60). Vaginal ACPAs positively correlated with *Streptococcus*; although Streptococci themselves do not express PADs, they can be citrullinated by neutrophil or bacterial PADs to activate ACPA-producing B cells(12). While vaginal detection of CRP and ACPAs have been previously reported(43, 58), this is the first report of vaginal RF to our knowledge. Vaginal ACPAs and RF co-occurred in most RA patients, yet no microbes and only four cytokines were simultaneously associated with ACPA and RF. These data suggest a complex RA vaginal autoimmune environment shaped primarily by CDAI and ACPA status that requires further experimental study to delineate shared and independent mechanisms.

There are several aspects of our study design that limit interpretation. Despite the broad demographic features captured in our cohort, we were limited to two clinics in a single medical center under the same providers. Care was taken to reduce over-fitting models due to our small sample size. Limitations also include the broad questionnaire which prevented further categorization of diet, restricted reporting of diabetes, autoimmune disease, and other medical conditions in the control group, and lack of viral and fungal components in analyses(10).

We describe correlations within the RA vaginal microbial and immunological landscape, although the directionality of these associations remains unknown. Despite overall shared CSTs and taxa between controls and RA, the functional capacity of microbes may be different given the unique RA immune profiles. *L. crispatus* strains from non-*Lactobacillus*-dominant communities express an additional glycosyltransferase compared to those in *Lactobacillus*-dominant communities(61), indicating the potential for acquired functions in different stress environments. Given the RA cohort did not exhibit increased urogenital symptoms nor prevalence of non-optimal CST IV, functional assays are necessary to verify the immunogenicity of bacteria in the context of RA. Likewise, longitudinal studies and larger cohorts are needed to assess infection risk and the impact of medication and RA flares on vaginal microbial composition. Despite vaginal immune signatures in bacterial vaginosis lacking systemic correlations(62), more work is needed to determine whether local immune dynamics instigate systemic changes or vice versa.

Ultimately, understanding the ties between the vaginal microbial and immune composition within RA will help inform patient care, monitoring, and risk assessment. Considering the shared impact of menopause-induced hormonal changes on the vaginal microbiome and RA pathogenesis(16), and the clear delineation of vaginal immune markers in post-menopausal RA and control patients, post-menopausal women may benefit the most from future vaginal intervention. If vaginal ACPA production contributes to disease onset, prophylactic treatment targeting specific taxa (e.g., *Streptococcus*) could aid in postponing or, ideally, preventing disease onset. If RA and/or RA medications are the driver of increased vaginal microbial alpha diversity, then reduction of inflammation or change in medication may ameliorate vaginal risk profiles in sexually active women(63). Addressing the vaginal environment as a potential site to suppress RA progression, disease activity, and RA-associated sequelae is key in improving the quality of life for women living with RA.

## Data Availability

All data produced in the present study are available upon reasonable request to the authors.

## ACKNOWLEDGEMENTS

We first want to thank the participants of the study. We also want to thank the Smith Clinic residents and faculty in the medicine clinic for facilitating sample collection and rheumatology attendings Dr. Rashmi Maganti and Dr. Shalini Jha for helping identify candidates at Smith and McNair Clinics. We additionally want to thank Sonia Mejia for translating the IRB to Spanish. We thank Zhongcheng Shi, Yuan Yao and Michael Nguyen from the BCM. Antibody-based Proteomics Core for their excellent technical assistance in performing the Luminex experiments, data preliminary analyses and QC, and project consultation. This work was supported in part by NCI Cancer Center Support Grant (P30CA125123) to the Antibody-based Proteomics Core. This work was supported by NIH T32 GM136554 and F31 AI167538 awards to M.E.M. Studies were supported by a Burroughs Wellcome Fund Next Gen Pregnancy Initiative (NGP10103), NIH R01 (DK128053), and NIH U19 (AI157981) awards to K.A.P.

## Author Contributions

M.E.M and K.A.P conceived and designed experiments. M.E.M, S.K.A, and K.A.P designed the study protocol and proposed and reviewed subsequent amendments. M.E.M, S.K.A, S.K, O.O, and L.L consented patients, recorded clinical metadata, and collected vaginal swabs. A.E, K.F, J.L, M.M, J.L, S.B, and Z.Z assessed RA patients and recorded clinical metadata. M.E.M, C.S, C.R, and M.B processed vaginal swabs. M.E.M, K.A.P, S.A, and S.H analyzed and interpreted results. M.E.M and K.A.P drafted the manuscript. K.A.P secured funding. All authors contributed to discussion and manuscript edits.

## REFERENCES

1. Myasoedova E, Crowson CS, Kremers HM, et al. Is the incidence of rheumatoid arthritis rising?: Results from Olmsted County, Minnesota, 1955-2007. Arthritis Rheum 2010;62.

2. Minichiello E, Semerano L, Boissier MC. Time trends in the incidence, prevalence, and severity of rheumatoid arthritis: A systematic literature review. Joint Bone Spine 2016;83.

3. van der Heijde D. Erosions versus joint space narrowing in rheumatoid arthritis: What do we know? Ann Rheum Dis 2011;70 Suppl 1.

4. Conforti A, Di Cola I, Pavlych V, et al. Beyond the joints, the extra-articular manifestations in rheumatoid arthritis. Autoimmunity reviews 2021;20.

5. Silman AJ,Pearson JE. Epidemiology and genetics of rheumatoid arthritis. Arthritis Res 2002;4 Suppl 3.

6. Mueller AL, Payandeh Z, Mohammadkhani N, et al. Recent advances in understanding the pathogenesis of rheumatoid arthritis: New treatment strategies. Cells 2021;10.

7. Wilson TM, Trent B, Kuhn KA, et al. Microbial influences of mucosal immunity in rheumatoid arthritis. Curr Rheumatol Rep 2020;22.

8. Hong M, Li Z, Liu H, et al. Fusobacterium nucleatum aggravates rheumatoid arthritis through fada-containing outer membrane vesicles. Cell Host Microbe 2023;31.

9. Maeda Y, Kurakawa T, Umemoto E, et al. Dysbiosis contributes to arthritis development via activation of autoreactive T cells in the intestine. *Arthritis & rheumatology (Hoboken*, N.J.) 2016;68.

10. Rojas M, Restrepo-Jiménez P, Monsalve DM, et al. Molecular mimicry and autoimmunity. J Autoimmun 2018;95.

11. Ercolini AM,Miller SD. Molecular mimics can induce novel self peptide-reactive CD4+ T cell clonotypes in autoimmune disease. Journal of immunology (Baltimore, Md.: 1950) 2007;179.

12. Brewer RC, Lanz TV, Hale CR, et al. Oral mucosal breaks trigger anti-citrullinated bacterial and human protein antibody responses in rheumatoid arthritis. Sci Transl Med 2023;15.

13. Scher JU, Ubeda C, Equinda M, et al. Periodontal disease and the oral microbiota in new-onset rheumatoid arthritis. Arthritis Rheum 2012;64.

14. Scher JU, Sczesnak A, Longman RS, et al. Expansion of intestinal Prevotella copri correlates with enhanced susceptibility to arthritis. eLife 2013;2.

15. Kvien TK, Uhlig T, Ødegård S, et al. Epidemiological aspects of rheumatoid arthritis: The sex ratio. Ann N Y Acad Sci 2006;1069.

16. Kanik KS,Wilder RL. Hormonal alterations in rheumatoid arthritis, including the effects of pregnancy. Rheum Dis Clin North Am 2000;26.

17. Hazes JM, Coulie PG, Geenen V, et al. Rheumatoid arthritis and pregnancy: Evolution of disease activity and pathophysiological considerations for drug use. Rheumatology (Oxford) 2011;50.

18. Shen L, Zhang W, Yuan Y, et al. Vaginal microecological characteristics of women in different physiological and pathological period. Frontiers in cellular and infection microbiology 2022;12.

19. Lan Y, Jin B, Zhang Y, et al. Vaginal microbiota, menopause, and the use of menopausal hormone therapy: A cross-sectional, pilot study in Chinese women. *Menopause (New York*, N.Y.) 2024;31.

20. Dabee S, Passmore JS, Heffron R, et al. The complex link between the female genital microbiota, genital infections, and inflammation. Infect Immun 2021;89.

21. Han Y, Liu Z, Chen T. Role of vaginal microbiota dysbiosis in gynecological diseases and the potential interventions. Front Microbiol 2021;12.

22. Lin CY, Lin CY, Yeh YM, et al. Severe preeclampsia is associated with a higher relative abundance of Prevotella bivia in the vaginal microbiota. Sci Rep 2020;10.

23. Davis NM, Proctor DM, Holmes SP, et al. Simple statistical identification and removal of contaminant sequences in marker-gene and metagenomics data. Microbiome 2018;6.

24. Bolyen E, Rideout JR, Dillon MR, et al. Reproducible, interactive, scalable and extensible microbiome data science using Qiime 2. Nat Biotechnol 2019;37.

25. McDonald D, Jiang Y, Balaban M, et al. Greengenes2 unifies microbial data in a single reference tree. Nat Biotechnol 2023;42.

26. Mandal S, Van Treuren W, White RA, et al. Analysis of composition of microbiomes: A novel method for studying microbial composition. Microb Ecol Health Dis 2015;26.

27. McMurdie PJ,Holmes S. Phyloseq: An R package for reproducible interactive analysis and graphics of microbiome census data. PLoS One 2013;8.

28. Wickham H. Ggplot2: Elegant graphics for data analysis: Springer-Verlag New York; 2016.

29. Itoh Y, Manaka M. Analysis of human vaginal secretions by SDS-polyacrylamide gel electrophoresis. Forensic Sci Int 1988;37.

30. Holm JB, France MT, Gajer P, et al. Integrating compositional and functional content to describe vaginal microbiomes in health and disease. Microbiome 2023;11.

31. France MT, Ma B, Gajer P, et al. VALENCIA: A nearest centroid classification method for vaginal microbial communities based on composition. Microbiome 2020;8.

32. Sun H, Guo Y, Wang H, et al. Gut commensal parabacteroides distasonis alleviates inflammatory arthritis. Gut 2023;72.

33. Unni R, Andreani NA, Vallier M, et al. Evolution of E. coli in a mouse model of inflammatory bowel disease leads to a disease-specific bacterial genotype and trade-offs with clinical relevance. Gut microbes 2023;15.

34. Zeng MY, Inohara N, Nuñez G. Mechanisms of inflammation-driven bacterial dysbiosis in the gut. Mucosal Immunol 2017;10.

35. Kumar BS, Suneetha P, Mohan A, et al. Comparison of disease activity score in 28 joints with ESR (DAS28), clinical disease activity index (CDAI), health assessment questionnaire disability index (HAQ-DI) & routine assessment of patient index data with 3 measures (RAPID3) for assessing disease activity in patients with rheumatoid arthritis at initial presentation. The Indian journal of medical research 2017;146.

36. Taylor PC, Chen YF, Pope J, et al. Patient disease trajectories in rheumatoid arthritis patients treated with Baricitinib 4-mg in four phase 3 clinical studies. Rheumatology and therapy 2023;10.

37. Fan J, Jiang T, He D. Advances in the implications of the gut microbiota on the treatment efficacy of disease-modifying anti-rheumatic drugs in rheumatoid arthritis. Front Immunol 2023;14.

38. Larsen JM. The immune response to Prevotella bacteria in chronic inflammatory disease. Immunology 2017;151.

39. Rioja I, Hughes FJ, Sharp CH, et al. Potential novel biomarkers of disease activity in rheumatoid arthritis patients: CXCL13, CCL23, transforming growth factor alpha, tumor necrosis factor receptor superfamily member 9, and macrophage colony-stimulating factor. Arthritis Rheum 2008;58.

40. Fukui S, Michitsuji T, Endo Y, et al. Distinct clinical outcomes based on multiple serum cytokine and chemokine profiles rather than autoantibody profiles and ultrasound findings in rheumatoid arthritis: A prospective ultrasound cohort study. RMD open 2025;11.

41. Ingegnoli F, Castelli R, Gualtierotti R. Rheumatoid factors: Clinical applications. Dis Markers 2013;35.

42. Dillon MC, Opris DC, Kopanczyk R, et al. Detection of homocysteine and C-reactive protein in the saliva of healthy adults: Comparison with blood levels. Biomark Insights 2010;5.

43. Di Naro E, Ghezzi F, Raio L, et al. C-reactive protein in vaginal fluid of patients with preterm premature rupture of membranes. Acta Obstet Gynecol Scand 2003;82.

44. Raposo B, Klareskog L, Robinson WH, et al. The peculiar features, diversity and impact of citrulline-reactive autoantibodies. Nature reviews. Rheumatology 2024;20.

45. Buchanan WW, Laurent RM. Rheumatoid arthritis: An example of ecological succession? Canadian bulletin of medical history = Bulletin canadien d’histoire de la medecine 1990;7.

46. Dhital R, Baer RJ, Bandoli G, et al. Cardiovascular events during pregnancy: Implications for adverse pregnancy outcomes in individuals with autoimmune and rheumatic diseases. The Journal of rheumatology 2025;52.

47. Skinner-Taylor CM, Perez-Barbosa L, Lujano-Negrete AY, et al. Sexual and reproductive health from the perspective of patients with autoimmune rheumatic diseases in Mexico: A qualitative study. BMJ open 2025;15.

48. Ostensen M, Brucato A, Carp H, et al. Pregnancy and reproduction in autoimmune rheumatic diseases. Rheumatology (Oxford) 2011;50.

49. Ling Z, Cheng Y, Gao J, et al. Alterations of the fecal and vaginal microbiomes in patients with systemic lupus erythematosus and their associations with immunological profiles. Front Immunol 2023;14.

50. Murphy EC, Frick IM. Gram-positive anaerobic cocci--commensals and opportunistic pathogens. FEMS Microbiol Rev 2013;37.

51. Chen J, Wright K, Davis JM, et al. An expansion of rare lineage intestinal microbes characterizes rheumatoid arthritis. Genome Med 2016;8.

52. Ilhan ZE, Łaniewski P, Tonachio A, et al. Members of Prevotella genus distinctively modulate innate immune and barrier functions in a human three-dimensional endometrial epithelial cell model. The Journal of infectious diseases 2020;222.

53. Nelson KG, Takahashi T, Bossert NL, et al. Epidermal growth factor replaces estrogen in the stimulation of female genital-tract growth and differentiation. Proc Natl Acad Sci U S A 1991;88.

54. Swanson CD, Akama-Garren EH, Stein EA, et al. Inhibition of epidermal growth factor receptor tyrosine kinase ameliorates collagen-induced arthritis. Journal of immunology (Baltimore, Md.: 1950) 2012;188.

55. Flytlie HA, Hvid M, Lindgreen E, et al. Expression of MDC/CCL22 and its receptor CCR4 in rheumatoid arthritis, psoriatic arthritis and osteoarthritis. Cytokine 2010;49.

56. T. Lebratti, Y. S. Lim, A. Cofie, et al. A sustained Type I IFN-neutrophil-IL-18 axis drives pathology during mucosal viral infection. Elife 2021;10.

57. Donmez HG, Cagan M, Fadiloglu E, et al. Is bacterial vaginosis associated with autoimmune antibody positivity? Cytopathology: official journal of the British Society for Clinical Cytology 2020;31.

58. Marcy D, Rothfuss H, Visser A, et al. Citrullinated peptide-specific ACPA are present in the female genital tract in premenopausal women with and without RA. Arthritis Rheumatology 2022;74. [abstract]

59. Cherrington B, Rothfuss H, Demoruelle K. The female cervicovaginal mucosa is a unique site for the production of autoantibodies associated with rheumatoid arthritis. Journal of Health Disparities Research and Practice 2018;12.

60. Derksen VFAM, Martinsson K, van Mourik AG, et al. Evidence of site-specific mucosal autoantibody secretion in rheumatoid arthritis. *Arthritis & rheumatology (Hoboken*, N.J.) 2025;77.

61. van der Veer C, Hertzberger RY, Bruisten SM, et al. Comparative genomics of human Lactobacillus crispatus isolates reveals genes for glycosylation and glycogen degradation: Implications for in vivo dominance of the vaginal microbiota. Microbiome 2019;7.

62. Hedges SR, Barrientes F, Desmond RA, et al. Local and systemic cytokine levels in relation to changes in vaginal flora. The Journal of infectious diseases 2006;193.

63. Zhang X, Zhang D, Jia H, et al. The oral and gut microbiomes are perturbed in rheumatoid arthritis and partly normalized after treatment. Nat Med 2015;21.

