## Supplemental Figures 1-4 for "A cross-sectional analysis of the vaginal microenvironment in rheumatoid arthritis"

### SUPPLEMENTAL MATERIAL

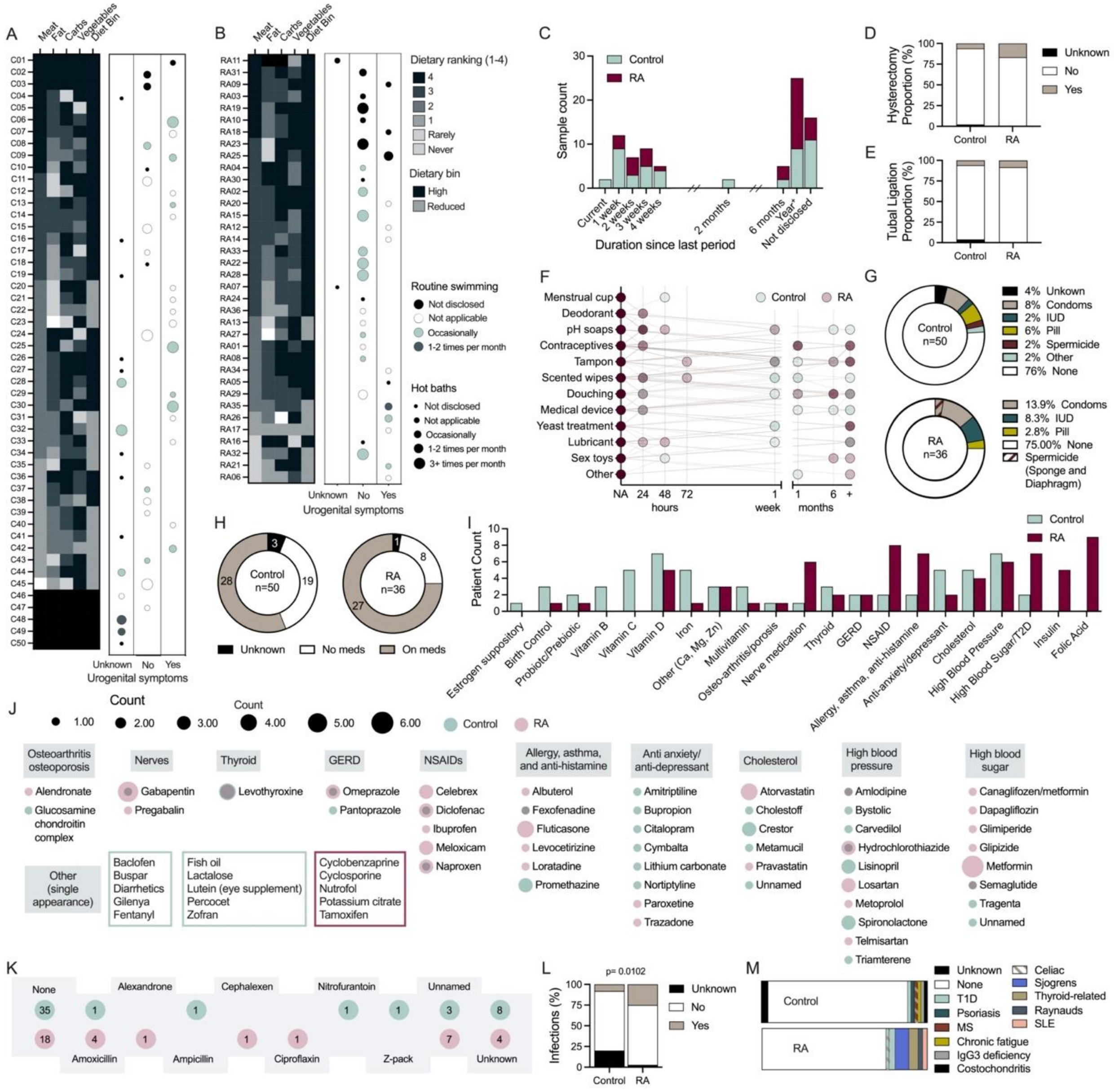


**Supplemental Figure 1. Medical information and recent practices of the study cohort.** Individual's diet breakdown and diet bin determination, and swimming and bath consistency stratified by presence of urogenital symptoms in (**A**) control and (**B**) RA groups. (**C**) Time since the start of the most recent period. Proportion of women with history of (**D**) hysterectomy or (**E**) tubal ligation. (**F**) Most recent use of products/items inserted into the vaginal canal; increasing intensity signifies multiplicity of use, and purple tones indicate overlap of control and RA use. (**G**) Contraceptive usage. (**H**) Prevalence and (**I**) types of medications or supplements in use. (**J**) Names of medications with overlapping use depicted by overlay of control (teal) and RA (pink) circles. (**K**) Antibiotics in use by control (bottom) and RA (top) individuals. (**L**) Proportion of women with recent infections. (**M**) Additional autoimmune diseases in each group. (D-E,L-M) Data were analyzed by Fisher's exact test. Significant *P*-values are shown.

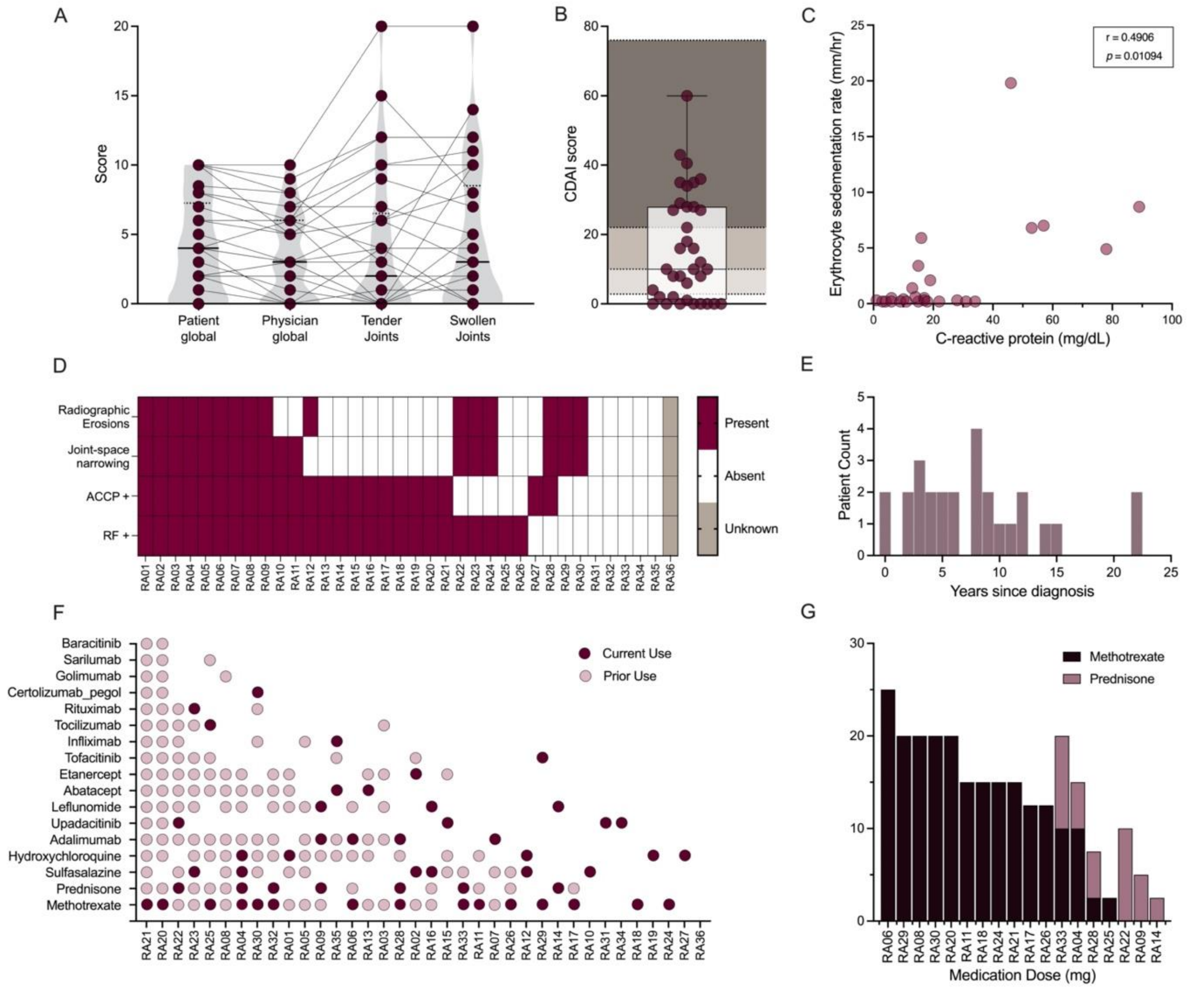


**Supplemental Figure 2. Clinical features within the RA cohort.** Disease severity was measured by compiling (**A**) patient and physician global scores, and tender and swollen joint counts to determine a (**B**) clinical disease activity index (CDAI) score. (**C**) Within individual erythrocyte sedimentation rate and serum C-reactive protein levels. (**D**) Presence of morphological symptoms and serum markers within individuals. (**E**) Duration of disease in years. (**F**) Medication history and (**G**) current dose prescribed for methotrexate and prednisone in current users (one dose not known). Symbols represent individuals with (A) repeats across clinical factor. (D,F) Each column represents one person. Data were analyzed by (C) Spearman correlation with *P*-value indicated.

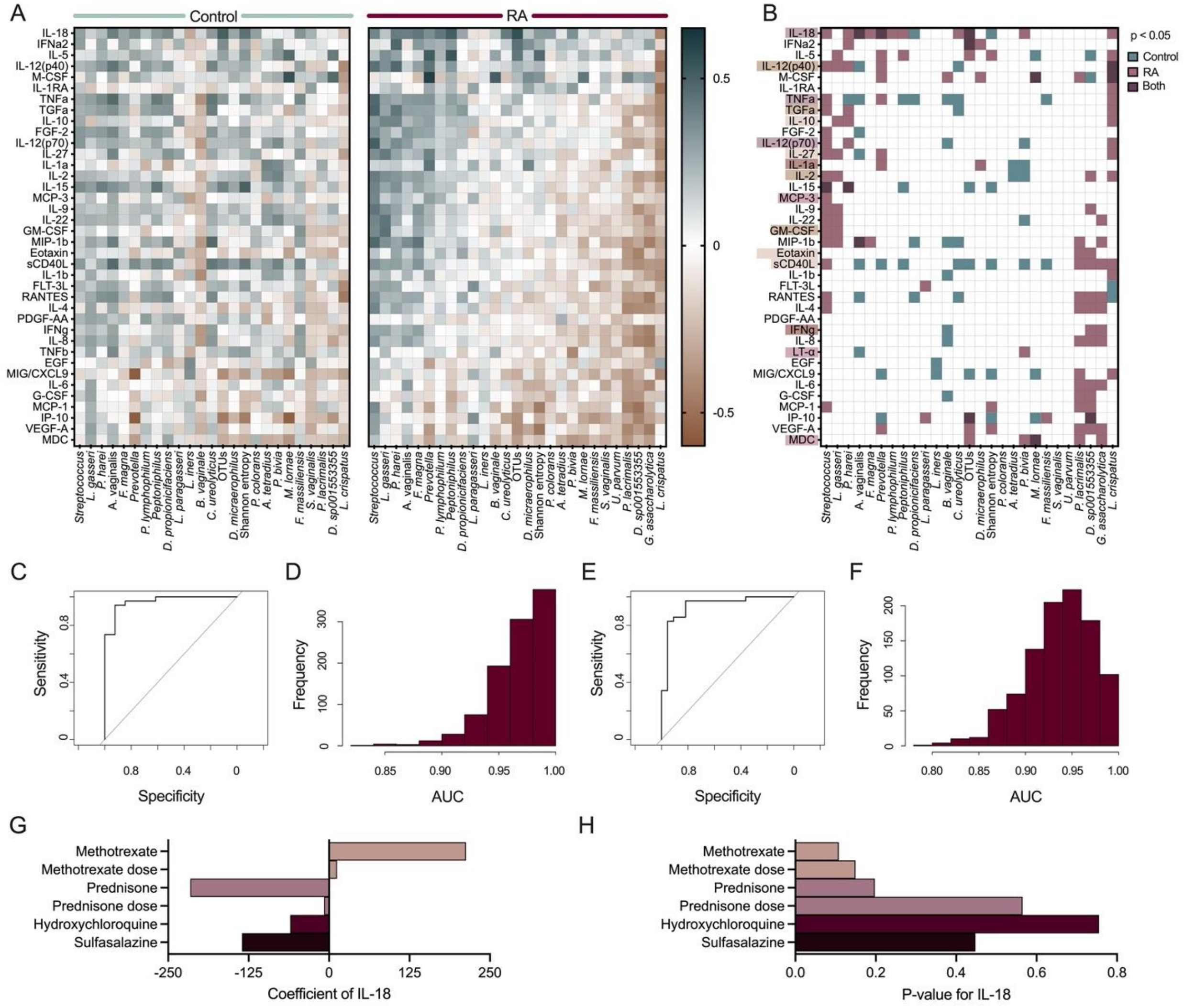


**Supplemental Figure 3. Correlations between immune factors and bacteria associated with RA or RA-related phenotypes.** (**A**) Matrices showing variable correlation coefficients between immune factors and significant microbes in control (left) and RA (right) groups. (**B**) Significant immune-microbe correlations established in the control (teal), RA (pink), or both (purple) vaginal environments. Immune markers are colored by corresponding RA metric associations shown in **Fig. 4**. RA was predicted by effects of *Prevotella*, *Peptoniphilus*, EGF and the interaction between sCD40L and diet, controlled for menopausal status and clinic. (**C**) AUC and (**D**) bootstrap distribution of RA prediction in a cohort of 34 RA and 13 controls. (**E**) AUC and (**F**) bootstrap distribution of RA prediction in a cohort of 35 RA and 22 unique controls. (**G**) Coefficient and (**H**) *P*-values of IL-18 association with medications. Data were analyzed by (A-B) Spearman correlation, (C-F) multiple logistic regression and (G-H) multiple linear regression. (C-F) Bootstrap sampling performed with 1000 iterations and seed set at 123

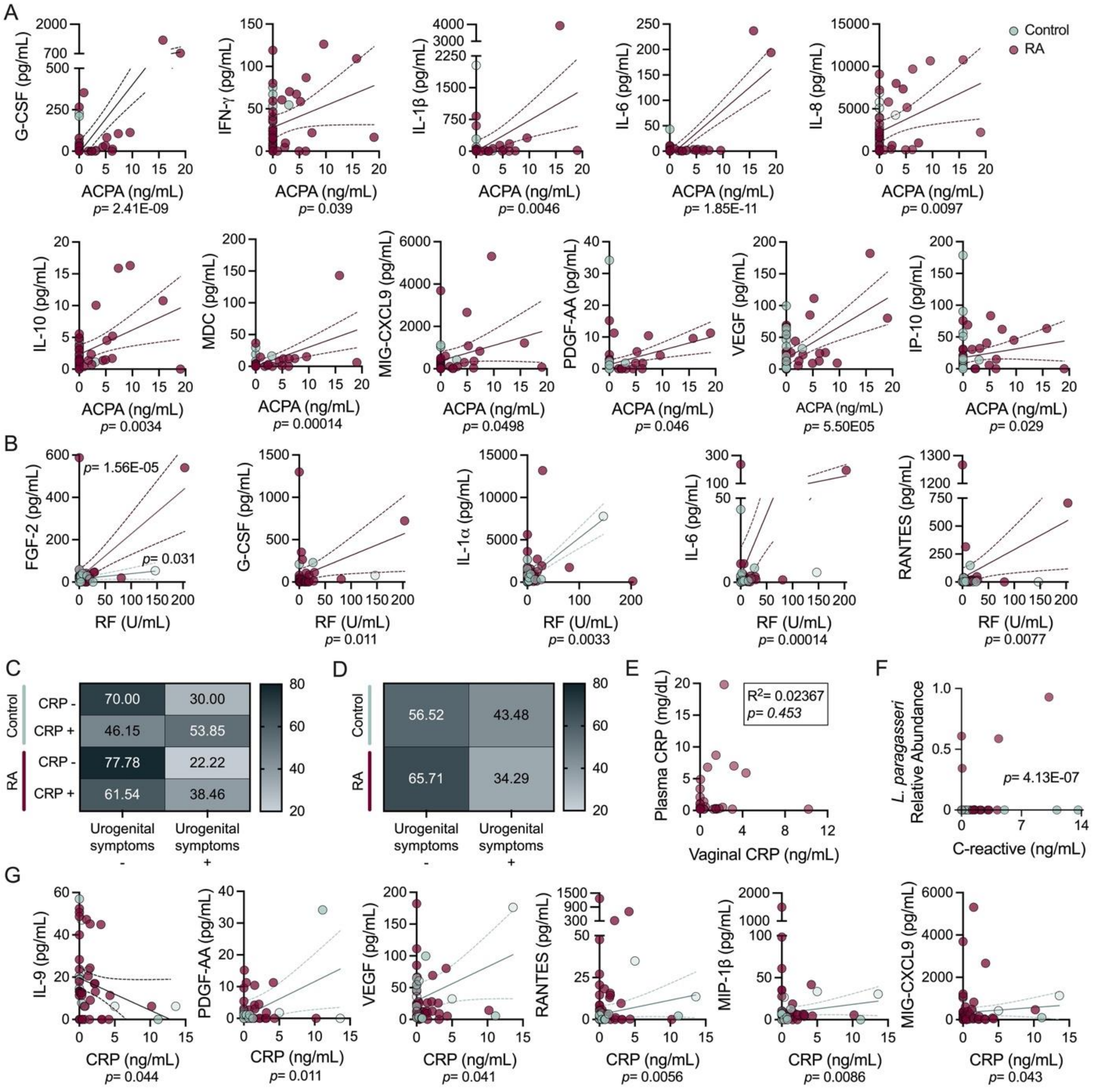


**Supplemental Figure 4. Vaginal levels of ACPAs, RF and CRP in correlation with cytokines and clinical history.** Correlations between immune factors and vaginal (**A**) ACPA and (**B**) RF. (**C**) Presence of urogenital symptoms according to vaginal CRP+/- status within the control and RA group or (**D**) regardless of CRP detection. (**E**) Correlation between vaginal concentrations of CRP and (**E**) plasma, (**F**) *L. paragasseri,* and (**G**) immune factors. Data were selected if (A-B,G) significant in multiple linear regression analysis and further assessed by Spearman correlations. Slope and 95% confidence interval are shown. Significant correlations in the RA group are dark red and light blue for the control group. (E) FGF-2 was significant for each group and IL-9 for both groups were combined. (A-B) Fisher's exact test and (E) multiple linear regression were also performed.
